# Safety and feasibility of lung biopsy in diagnosis of acute respiratory distress syndrome: protocol for a systematic review and meta-analysis

**DOI:** 10.1101/2020.10.21.20201244

**Authors:** Yosuke Fukuda, Hiroshi Sugimoto, Yoshie Yamada, Hiroyuki Ito, Takeshi Tanaka, Takuo Yoshida, Satoshi Okamori, Koichi Ando, Yohei Okada

## Abstract

**Introduction:** Acute respiratory distress syndrome (ARDS) is a type of acute respiratory failure characterized by non-cardiac pulmonary edema caused by various underlying conditions. ARDS is often pathologically characterized by diffuse alveolar damage (DAD), and its pathological findings have been reported to be associated with prognosis, although the adverse effects of lung biopsies to obtain pathological findings are still unclear. The purpose of this systematic review and meta-analysis is to reveal the safety and feasibility of lung biopsy in the diagnosis of ARDS.

**Methods and analysis:** We will include studies that were published in MEDLINE and Cochrane Central Register of Controlled Trials until June 1, 2020. We will include the reports for critically ill patients in an intensive care unit or emergency department who undergo lung biopsy and require a mechanical ventilation. Two review authors will independently scan titles and abstracts of all identified studies. Furthermore, these two authors will read and assess the full text of study reports to identify trials that appeared broadly to address the subject of the review. We will perform a risk of bias assessment using the McMaster Quality Assessment Scale of Harms.

**Ethics and dissemination:** This study will be based on the published data, therefore, it does not require ethical approval. The final results of the study will be published in a peer-reviewed journal.

**UMIN registration number:** UMIN000040650

**Strengths and limitations of this study:**

- This protocol complies with the Reporting Items for Systematic Review and Meta-Analysis Protocol guidelines.
- This systematic review and meta-analysis will assess the safety and feasibility of lung biopsy in patients with ARDS.
- We will evaluate the risk of bias and report according to the McMaster Quality Assessment Scale of Harms.
- This review will include the reports about only adult patients with acute respiratory failure.
- We will plan to exclude non-English databases in this study.

## INTRODUCTION

Acute respiratory distress syndrome (ARDS) is a type of acute respiratory failure characterized by non-cardiac pulmonary edema caused by various underlying conditions.[1] Many conditions, including pneumonia, sepsis, and trauma, have been considered as a trigger of ARDS.[2] In intensive care units (ICUs), the incidence of ARDS is 10.4% over 4 weeks.[2] Although the mortality rate of ARDS has declined in recent years with advances in understanding the disease and treatment of the disease, it remains high at about 30%.[2, 3]

Although ARDS is a group of diseases characterized by pathologically diffuse alveolar damage (DAD),[5] a previous study reported that only 45% of patients with ARDS who met Berlin criteria had DAD based on autopsy cases.[6] However, it was reported that ARDS, which is pathologically DAD, had a poor prognosis.[7] The mainstay of the management of ARDS is the treatment of the triggers and supportive mechanical ventilation,[5] therefore, it is essential to confirm the ARDS diagnosis pathologically and rule out other conditions that have specific treatment,[5] and the results of lung biopsies may be used to predict the prognoses of patients with ARDS.

There are some procedures to perform lung biopsy to pathologically validate ARDS, including transbronchial lung biopsy (TBLB), cryobiopsy, and surgical lung biopsy (SLB).[8] Although they are useful approaches for the pathological diagnosis, various complications may occur during and after the procedures. Previously, several studies showed that 18%-59% of patients who underwent SLB experienced biopsy-related complications.[9-11] Major complications include pneumothorax, prolonged air leak, bleeding, and infection.[8] However, little is known on the prevalence in detail, and the harmfulness of lung biopsy for ARDS remains controversial. Therefore, we conducted a systematic review and meta-analysis to examine the incidence of adverse events or concerns of the safety of lung biopsy for patients with acute respiratory failure, including ARDS.

The objectives of our systematic review are to investigate the incidence of adverse events or concerns of the safety of lung biopsy for adult patients with acute respiratory failure including ARDS in an ICU or emergency department (ED).

## METHODS AND ANALYSIS

### Eligibility criteria

We will include all the reports in which critically ill patients in an ICU or ED undergo lung biopsy and require mechanical ventilation. This systematic review will include randomized controlled trials and observational studies (cross-sectional studies, prospective cohort studies, and retrospective cohort studies). We will exclude case reports, case-control studies, and review articles.

### Participant eligibility

This systematic review will target participants as follows;

1. Adult patients who are 16 years or older.
2. Critically ill patients in an ICU or ED setting.
3. Requiring mechanical ventilation for acute respiratory failure.
4. Undergo lung biopsy

We define TBLB, cryobiopsy, and SLB (video-assisted thoracoscopic surgery or open lung biopsy) as lung biopsy procedures. All patients who meet the above criteria will undergo a lung biopsy procedure under mechanical ventilation.

### Outcome measures and analysis

Primary outcome measures are biopsy-related death, respiratory failure, cardiac complication, bleeding, and other major complication in patients with lung biopsy. Secondary outcomes are pneumothorax, infection, cost, human cost, and other minor complications to be measured in those with lung biopsy. We set these outcomes according to the British Thoracic Society guidelines for diagnostic flexible bronchoscopy in adults.[12]

### Electronic searches

A systematic search of the literature will be conducted according to the Preferred Reporting Items for Systematic Reviews and Meta-Analysis Statement.[13] In electronic searches, we will consult with librarians to conduct systematic searches. There is no publication year or publication status restrictions. We will search MEDLINE and Cochrane Central Register of Controlled Trials (CENTRAL).

### Search strategy

Our initial search syntax for MEDLINE and CENTRAL is presented in Tables 1 and 2.

**Table 1.** Search strategy for MEDLINE.

| Search Number | Query |
| --- | --- |
| #1 | Respiratory Distress Syndrome, Adult [mh] |
| #2 | Acute lung injury [mh] |
| #3 | ALI [tiab] OR ARDS [tiab] |
| #4 | Acute [tiab] AND (lung injur* [tiab] OR respiratory distress [tiab] OR respiratory failure[tiab]) |
| #5 | (Severe [tiab] OR critical*[tiab]) AND (respiratory[tiab] OR hypox* [tiab]) |
| #6 | #1 OR #2 OR #3 OR #4 OR #5 |
| #7 | biopsy[mh] AND lung[mh] |
| #8 | (cryosurger*[tiab] OR cryobiopsy[tiab] OR biopsy[tiab]) AND lung[tiab] |
| #9 | bronchoscopy[mh] |
| #10 | Thoracic Surgery, Video-Assisted[mh] |
| #11 | #7 OR #8 OR #9 OR #10 |
| #12 | #6 AND #11 |
| #13 | animals[mh] NOT human[mh] |
| #14 | #12 NOT #13 |

**Table 2.** Search strategy for Cochrane Central Register of Controlled Trials (CENTRAL)

| ID | Search |
| --- | --- |
| #1 | [mh "Respiratory Distress Syndrome, Adult"] |
| #2 | [mh "Acute lung injury"] |
| #3 | ALI:ti,ab OR ARDS:ti,ab |
| #4 | Acute:ti,ab |
| #5 | lung NEXT injur*:ti,ab |
| #6 | "respiratory distress":ti,ab |
| #7 | "respiratory failure":ti,ab |
| #8 | #4 AND (#5 OR #6 OR #7) |
| #9 | (Severe:ti,ab OR critical*:ti,ab) AND (respiratory:ti,ab OR hypox*:ti,ab) |
| #10 | #1 OR #2 OR #3 OR #8 OR #9 |
| #11 | [mh biopsy] AND [mh lung] |
| #12 | (cryosurger*:ti,ab OR cryobiopsy:ti,ab OR biopsy:ti,ab) AND lung:ti,ab |
| #13 | [mh bronchoscopy] |
| #14 | [mh "Thoracic Surgery, Video-Assisted"] |
| #15 | #11 OR #12 OR #13 OR #14 |
| #16 | #10 AND #15 |
| #17 | [mh animals] NOT [mh human] |
| #18 | #16 NOT #17 |

### Data collection and analyses

We will implement the search strategies and import all references to EndNote X9 (Clarivate Analytics, Tokyo, Japan). Then, the results from the different electronic databases will be combined in EndNote library, and we will remove duplicates. Two authors independently select data from the studies using standardized data forms, including the following information;

1. Study characteristics: author, year of publication, country, design, sample size, clinical settings, number studied, and funding source.
2. Population characteristics: inclusion/exclusion criteria, number of drop-outs with reason, and patient demographics such as age and sex.
3. Intervention of interest: which technique is used in the lung biopsy, when it is performed, where it is performed, and who performs it.
4. Outcomes: We will search for details and frequency of adverse events in lung biopsy.

Two review authors will independently scan titles and abstracts of all identified studies. Then these two authors will read the full text of study reports and assess to identify trials that appeared broadly to address the subject of the review. Both authors scrutinize the full text of these articles for eligibility. When there is disagreement between the authors on the inclusion and exclusion criteria, we will discuss it and reach a consensus decision.

### Assessment of methodological quality

Two investigators will independently evaluate the risk of bias and report according to the McMaster Quality Assessment Scale of Harms (McHarm).[14] A statistical assessment of publication bias will not be performed. Therefore, we look at the number of ongoing and unpublished studies for the assessment of publication bias.

### Data Synthesis

In this study, we will perform a random-effects meta-analysis based on the DerSimonian and Laird method.[15] Forest plots will be generated with its 95% confidence intervals (CIs). When integrating extracted data from single-arm primary studies, the pooled effect size will be expressed as the population ratio and its 95% CI. When integrating extracted data from primary studies with arms of the comparison or control groups, the pooled effect size will be expressed as the risk ratio (RR) or odds ratio (OR); if the 95% CI crosses the invalid line (i.e., the OR or RR was 1), the results are considered to be insignificant. These analyses will be conducted only if the extracted data allow them. All analyses will be performed using SAS, STATA, R software, or Review Manager 5.3 (Cochrane Collaboration, London, United Kingdom). Finally, we will prepare a summary of findings table detailing the studies of concern (patient population, lung biopsy procedure, actual number and frequency death, serious complications, complications requiring additional treatment, prolonged duration of treatment, minor complications, costs, and human costs). If we determine that the data cannot be merged because of substantial heterogeneity, we will not perform a meta-analysis.

### Investigation of heterogeneity

Heterogeneity will be quantified by using the *I*^2^ statistical method. We will perform the subgroup analyses on the following groups if available; different patients characteristics, definition of the patients, index test, and reference standard.

### Sensitivity analysis

We will assess the robustness by excluding the studies with a high risk of bias.

### Patient and public involvement

Patient and public involvement is not required for this systematic review.

## ETHICS AND DISSEMINATION

This study will not need ethics approval because we will use only published data. This systematic review and meta-analysis will be published in a peer-reviewed journal and presented at a scientific conference relevant to this field.

## Supporting information

Supplemental Table 1. PRISMA-P check list

## Data Availability

Data sharing is not applicable to this article as no new data were created or analyzed in this study.

## Acknowledgements

We thank Editage (www.editage.jp) for English Language editing.

## Authors’ contributions

YF, HS and YO conceived the original idea for this systematic review. YF and HS drafted the manuscript. YY, HI, TT, TY, SO, KA and YO revised the manuscript. All authors have read and approved the final manuscript.

## Patient and public involvement

This study will be based on the published data. Patients and public will not be involved in the design and conduct of the study, choice of outcome measures, recruitment to the study, and dissemination of the study.

